# Barriers and facilitators for digital health medical device registration in the UK: A scoping review

**DOI:** 10.1101/2025.04.13.25325766

**Authors:** Madison Milne-Ives, Katie Bounsall, Ananya Ananthakrishnan, Rosiered Brownson-Smith, Cen Cong, Camille Carroll, Edward Meinert

## Abstract

**Background:** Digital technology within healthcare has dramatically expanded in recent years, particularly with the growth of the use of medical or health-related apps. With this expansion comes the need for regulation and certification; however, the regulation process has many barriers. No review has examined the barriers and facilitators for stakeholders who aim to register software as a medical device.

**Methods:** The review was structured using the PRISMA extension for Scoping Reviews and PICOS framework. Studies were retrieved from five databases: PubMed, Embase, Cumulative Index to Nursing and Allied Health Literature, Web of Science, and Scopus. A descriptive analysis provided an overview of the barriers and facilitators to the registration of digital health solutions as medical devices in the UK.

**Results:** A total of 9 studies met the inclusion criteria and were included in the analysis. Of the 9 studies, the most common were perspective papers (n=4), with the remaining consisting of qualitative research (n=1), case studies (n=2), a development framework (n=1) and a debate paper (n=1). 4 of the 9 studies discussed any digital technologies, 3 discussed mental health apps, and the remaining 2 studies discussed a range of open-source hardware, mobile health care technologies and mental health apps. The analysis identified a range of barriers, including complexity, lack of clarity - particularly in what constituted a medical device - and a lack of tailoring of the process to specific characteristics of digital health technologies. The process of registration was described as confusing and complex. Analysis of facilitators highlighted several guidance documentations to support registration and a tiered approach to registration based on the intended use and risk level. Some studies highlighted potential ways to improve the process, including templates and examples approved by notified bodies, medical device certification training, and greater evidence-generation flexibility.

**Conclusions:** This review highlights the importance of clarity and specificity in definitions and guidance for registering and regulating digital health technologies. It also opens avenues for further discussion on levels of influence of registration barriers on patients, healthcare professionals and other end users to reflect user needs in registration. Further exploration of registrations with other regulatory jurisdictions and real-life cases of registration are still required to extend the understanding of barriers to registration beyond the UK context.

**Author Summary:** This review explored the challenges to stakeholders in the registration of digital health technologies as medical devices in the UK. We identified a few key challenges: complex registration regulations, unclear definitions of medical devices and clinical validation, and a lack of clinical evidence requirements for digital health technologies. These challenges would make it difficult for stakeholders to determine if their software qualifies as a medical device and how to demonstrate its safety and efficacy. High costs associated with registration and complex clinical validation requirements are particularly challenging for startups and SMEs, given their limited resources and experience. Existing guidance and tiered classification of medical devices facilitate the registration process to some extent, but not sufficiently for stakeholders without relevant expertise. To facilitate the registration process, we encourage templates and examples from approved bodies for medical devices, as well as tailored guidelines of device registration for different types of digital health technologies.

## Introduction

### Background

The expanding influence of digital technology in human activity has been called the “Fourth Industrial Revolution”, and this is no different in the field of healthcare ^1^. Digital health technology has increasingly become a policy focus to improve the efficiency and quality of healthcare delivery. Within the UK’s National Health Service (NHS), a shift from analogue to digital is underway, with a focus of innovation ^2^. Technologies or digital tools such as wearables and mobile health applications are crucial to modernise healthcare services ^3^. To ensure that this technology is safe and beneficial for patients, there is a need for regulation - just as in the pharmaceutical industry. Although there is a movement toward increasing regulation of digital health technologies, existing regulatory systems and legislation are still being adapted to make clear their applications for software ^4^. In the UK, the Medicines and Healthcare products Regulatory Authority (MHRA) is responsible for regulating medical devices, including software ^5^. To ensure proper certification, developers of digital health technologies must navigate a complex and often confusing medical device registration process. This review compiles barriers and facilitators that have been identified within this process in the UK to provide an overview of areas where the process could be improved to facilitate medical device registration for digital health technologies.

In 2012, there were estimated to be at least 40,000 medical and health-related apps available ^6^; by 2021, this had increased to almost 100,000 and continues to grow ^7^. In the UK, digital technology is a key element of both the NHS Long Term Plan in 2019 and the new vision in spring 2025 ^8,9^ to improve service delivery and empower patients to manage their own health. Digital health technologies have patient-facing, point-of-care, and backend applications ^10^ and can serve a variety of purposes, including prevention of disease, behaviour change support, diagnosing and monitoring diseases, storing and analysing medical or well-being data, and clinical decision support/triage tools, among others ^11^.

The relative recency and rapid development of digital technology in healthcare means that regulatory processes are still catching up to the specifics of digital health technologies ^12^, which has the potential to create ambiguity for developers. The abundance of available digital health technologies can also make it difficult for patients and healthcare providers to identify safe and high quality technologies ^4,13^. Previous studies have found that many mobile health apps that would be classified as medical devices have not been certified as such, and thus avoid regulation ^5^. On the other hand, there is little regulation for mobile health apps that are not considered medical devices ^14^. Regulation is important to ensure that digital health technology is safe, secure, and effective and to help users access the best and most appropriate tools ^6^.

Medical device regulation guidance is constantly being reviewed and updated. The MHRA revised its roadmap to the implementation of a medical devices regulatory reform in December 2024 ^9^ and updated its guidance on software and artificial intelligence as a medical device on 3rd February 2025 ^15^. To inform future regulations, the MHRA consulted with key stakeholders (including healthcare professionals and institutions, manufacturers, members of the public/patients, trade associations, and SMEs) on the proposed changes ^16^. The current process for registration and regulation has a range of different contributors. It is a legal requirement within the UK to be regulated under the Medical Devices Regulations 2002 (UK MDR 2002) which includes the directive 93/42/EEC on medical devices (EU MDD). In the EU, conformity assessments are conducted by notified bodies ^17^. Despite the MHRA being responsible for the regulation of medical devices (including software), manufacturers are also required to achieve the UK Conformity Assessed (UKCA) standard, which incorporates CE Marking (Conformité Européene), and specific to digital health tools, Digital Technology Assessment Criteria (DTAC). Since 31st December 2024, the CE Marking is no longer valid and must be replaced with UKCA. The UK’s National Institute for Health and Care Excellence (NICE) published an evidence standards framework (ESF) in 2022. It provides standards for the evidence that should be applied to digital health technologies to demonstrate their value.Although these standards are not mandatory from a regulatory perspective, the ESF describes the different levels and types of evidence a digital health technology will need in order to be commissioned within the UK. It is a standalone assessment and does not contribute towards MHRA, UKCA or DTAC applications.

Although there is an existing body of literature examining challenges to the regulation of digital health technologies in general ^4,18–20^, no previous reviews were found that specifically examined the issues faced by stakeholders who are aiming to register their software as a medical device. This is an important gap to address to support continued growth and innovation in digital health technology while ensuring high-quality software, as developers must register their products with the MHRA before they can be made available in UK markets. This scoping review aimed to provide an overview of the barriers and facilitators involved in the process of medical device registration for digital health solutions in the UK.

## Materials and Methods

### Scope

The review was structured using the Preferred Reporting Items for Systematic Reviews and Meta-Analyses Extension for Scoping Reviews (PRISMA-ScR; Supplemental Material A) guidance ^21^ and the Population, Intervention, Comparator, Outcome, and Studies (PICOS) framework (Table 1).

**Table 1.**
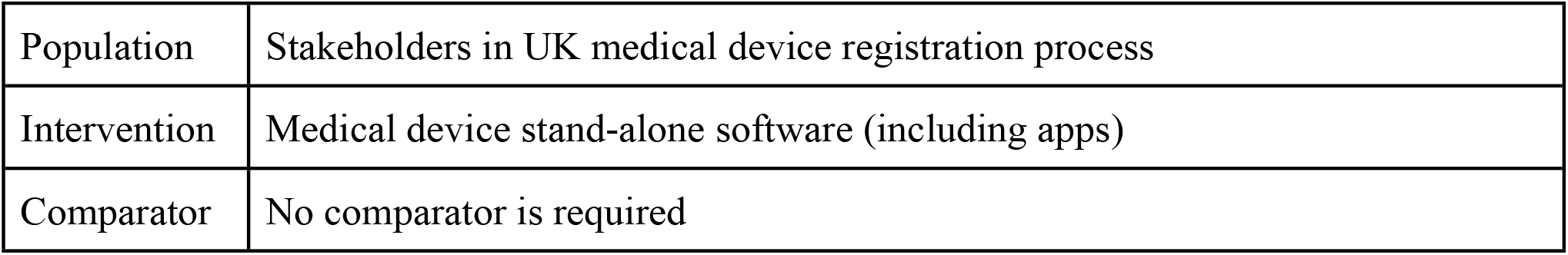
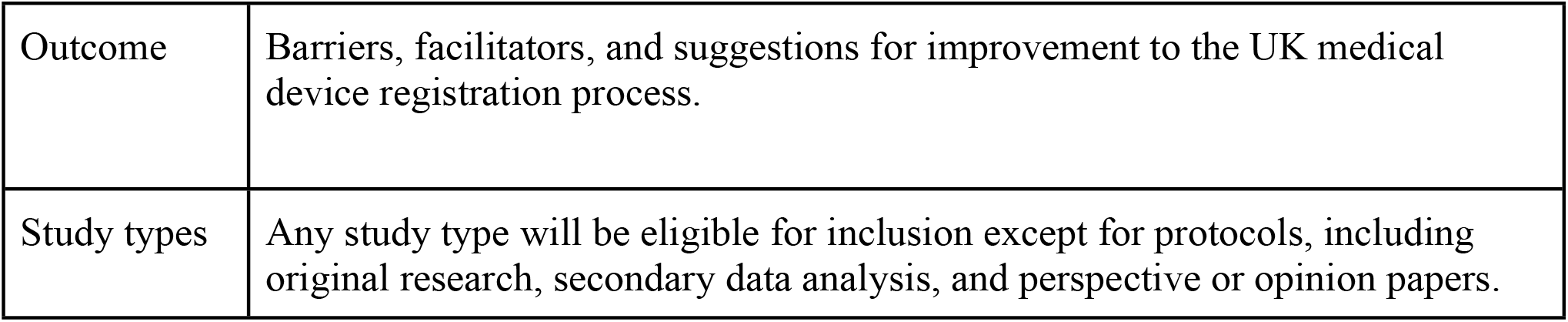
PICOS framework.

### Search strategy

Six databases (PubMed, Embase, Cumulative Index to Nursing and Allied Health Literature (CINAHL), Web of Science, Scopus, and Google Scholar were selected to be searched to provide a wide coverage of sources relating to health and information and computing technology. The search was conducted in October 2024. The search string (Table 2) was developed based on a preliminary review of the literature to identify key terms and structured as: digital health software (MeSH OR Keywords) AND regulatory process (MeSH OR Keywords). Full search strings and references retrieved are included in Supplemental Material B. The search conducted in Google Scholar identified 1,520 references, but it was not possible to export them all together, and an attempt to download them individually resulted in Google Scholar limiting the amount of references that could be exported to about 100. Due to time constraints, this meant that Google Scholar had to be removed from the search.

**Table 2.** Search string.

| Category | MeSH | Keywords |
| --- | --- | --- |
| Software applications | Cell Phone OR Telemedicine OR Mobile Applications | “Smartphone” OR “mobile phone” OR mHealth OR “mobile health” OR “eHealth” OR “digital health” OR “chatbot” OR “wearable” OR “telehealth” OR “mobile app” OR “smartphone app” OR “medical informatics application” OR “web app” OR “internet-based application” OR “software applications” OR “internet of things” |
| Regulatory process | Government Regulation | “MDR” OR “medical device registration” OR “medical device regulation” OR “UKCA” OR “CE mark” OR “MHRA” OR “regulatory” OR “class I” OR “class II” OR “class III” OR “medical device” OR “certification” OR “DTAC” |

### Screening and article selection

The citation management software EndNote X9 was used to store exported references. After automated duplicate removal, one author (MMI) used the EndNote X9 search function to conduct an initial keyword-based screening of references (Supplemental Material 3) and manually screened the remaining titles and abstracts (MMI). The screening was conducted from 28 October to 10 November 2022. Two authors (MMI & KB) independently screened the full-texts of the articles and then resolved any discrepancies through consensus to determine final eligibility. Screening was deliberately inclusive at the title and abstract stage, to avoid eliminating studies where barriers or challenges were mentioned in the body of the text but were not the main focus of the paper.

### Eligibility criteria

A broad collection of search terms was used initially to reduce the likelihood that relevant articles were missed in the search. The scope of the articles was narrowed in the screening stage based on the following inclusion and exclusion criteria.

#### Inclusion criteria

Studies were eligible for inclusion if they discussed at least one barrier or facilitator to the UK medical device registration process. To capture a larger body of literature and avoid the effect of Brexit on recent policy, studies discussing the UK MHRA, EU MDR, or CE marking medical device registration processes were included. Interventions addressed could be any type of digital software with a medical purpose that would make it potentially eligible for medical device registration. Beyond that, there were no limits on the type of software or on the aspect of healthcare they were addressing.

#### Exclusion criteria

The review aimed to focus on contemporary processes of medical device registration that reflect the current landscape of digital health technologies. Studies published before 2008 were excluded; this cut-off was selected as it is when smartphones became available and marks the shift into an age where mobile and other digital devices were ubiquitous amongst the general population. Studies published in languages other than English were also excluded, as the authors did not have the ability to read them. Studies that conducted evaluations of interventions without discussing their regulation were excluded, as were studies that focused on challenges to software as medical device regulation in general (rather than the specific medical device registration process). Studies that did not consider UK (or constituent nation) medical device registration processes specifically - either fully or as one of a number of jurisdictions examined - were also excluded.

### Data extraction

Data was extracted from the included studies into a pre-developed table by two reviewers (Table 3).

**Table 3.** Data extraction table.

| Article Information | Data to be extracted |
| --- | --- |
| General study information |  |
|  | Year of publication |
|  | Study/paper type |
|  | Sample size |
|  | Sample details |
| Intervention |  |
|  | Type of software medical device |
|  | Regulation type |
| Evaluation |  |
|  | Barriers/challenges to medical device registration |
|  | Facilitators to medical device registration |

### Data analysis and synthesis

One author conducted a descriptive analysis and narrative synthesis of the data extracted from the studies to synthesise the barriers and facilitators experienced in the process of registering digital health solutions as medical devices in the UK.

## Results

### Included studies

The database search retrieved 12,087 references (Supplemental Material 2). 3,141 duplicates were removed using the EndNote X9 software and a screening using the EndNote X9 keyword search tool resulted in 191 studies for title and abstract review (Supplemental Material 3).

One reviewer screened the titles and abstracts of the 191 studies using Rayyan software ^22^ and two reviewers screened the remaining 57 full-text articles that were identified as potentially eligible. Figure 1 provides an overview of this process with reasons for exclusion at full-text stage.

**Figure 1.**
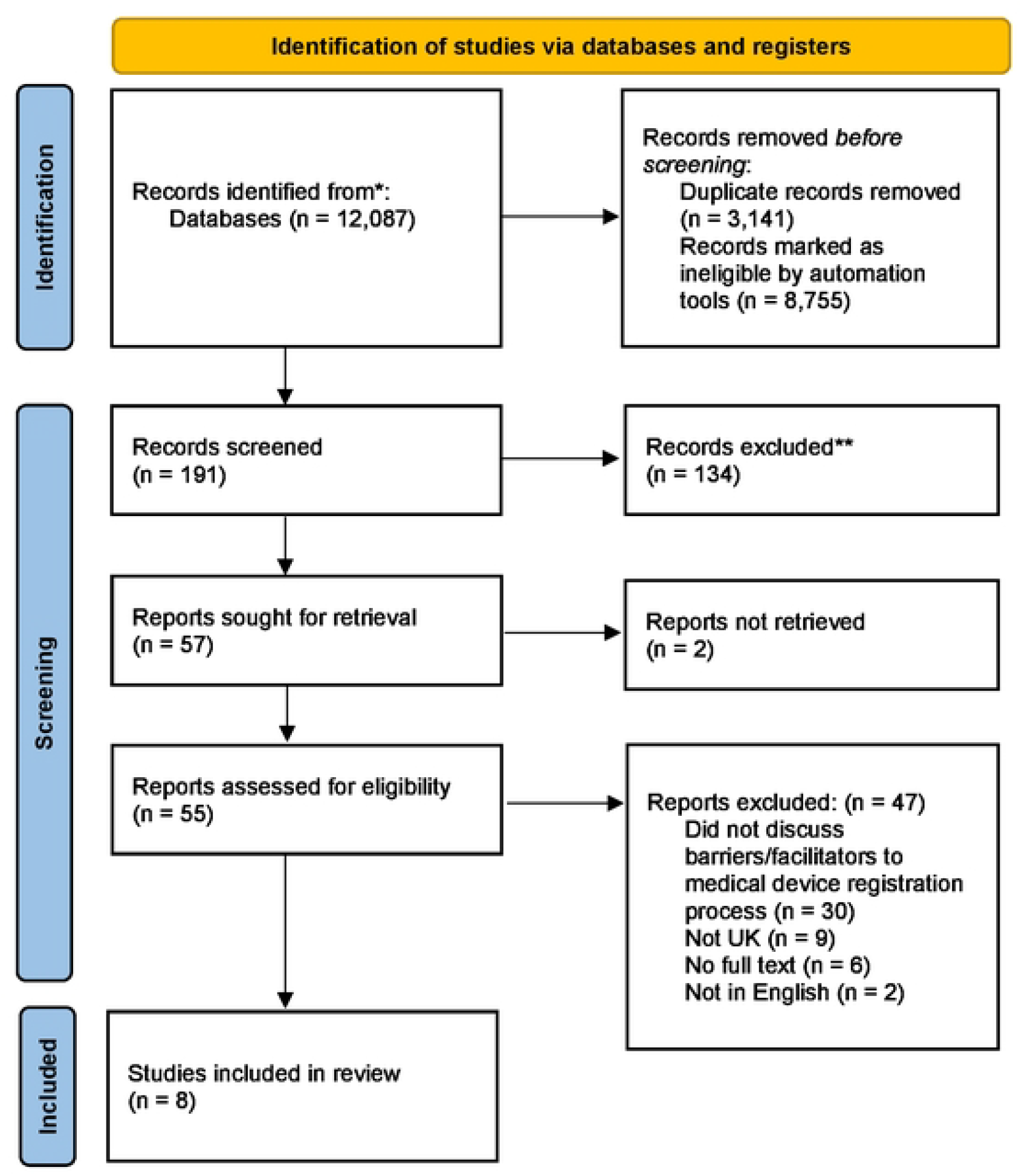
Preferred Reporting Items for Systematic Reviews and Meta-Analyses (PRISMA) flow diagram

### Study characteristics

The main characteristics of the studies and the interventions they consider are reported in Table 4.

**Table 4.**
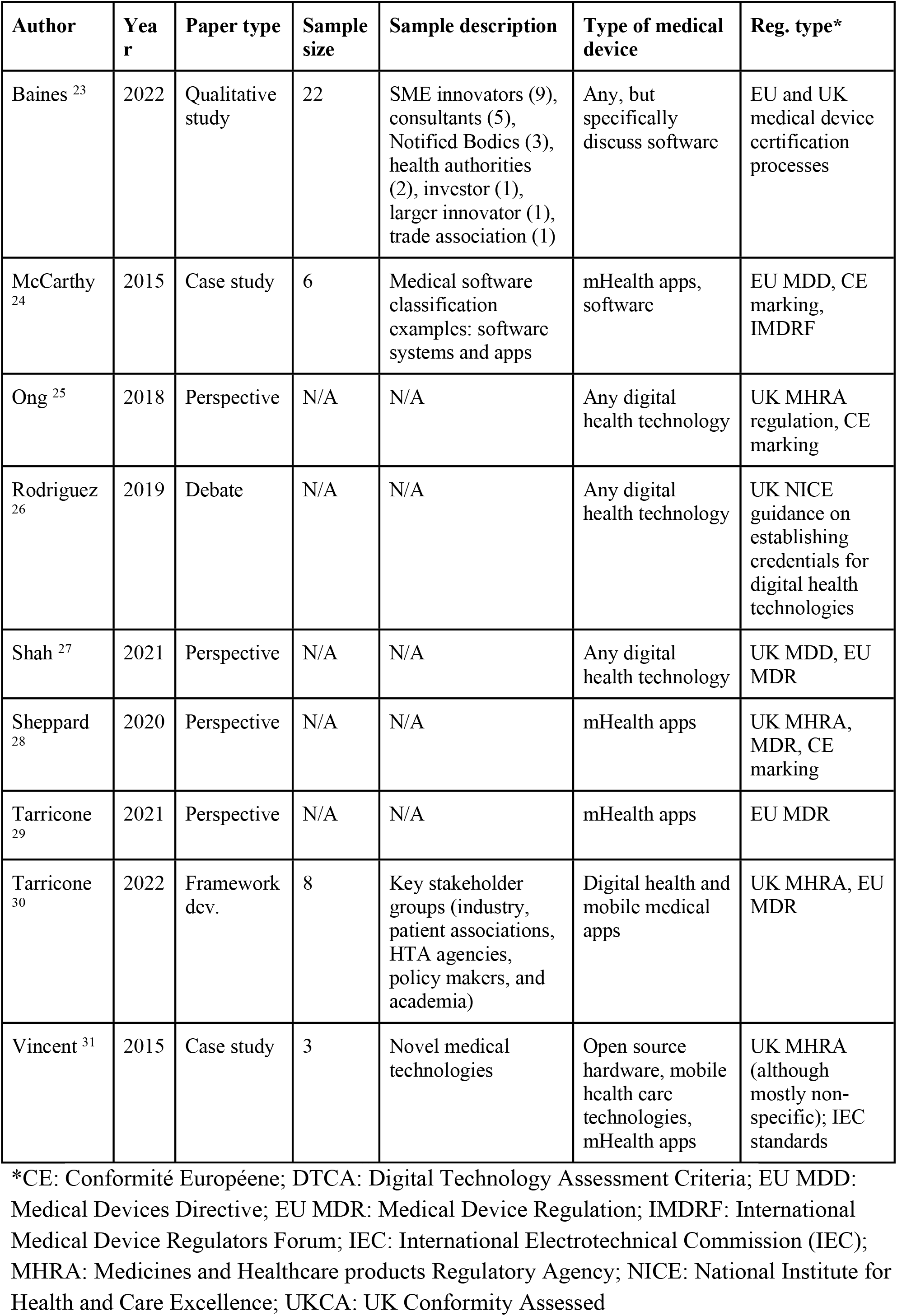
Summary of study characteristics for included articles.

#### Barriers to medical device registration for software

The analysis of the articles identified several barriers to the UK’s medical device registration process; these are summarised in Table 5. Overall, stakeholders considered the regulatory rules and accreditation process confusing and complex ^23^The time, costs and expertise required to complete the process and provide sufficient evidence of clinical safety and efficacy can be a barrier for all companies but is particularly relevant ^23,28^ for startups and small and medium-sized enterprises (SMEs), who may lack the experience or in-house templates and examples needed to facilitate independent completion of the registration process ^23,24^. Notified bodies are responsible for assessing medical devices for regulatory approval but they are not allowed to provide direct guidance or consultation to companies during the regulatory process to remain independent This caused frustration for companies and increased the difficulty of completing the registration process ^23^. Other common identified barriers concerned definitions of what a medical device is and clinical validation requirements ^24,25,28,31^.

**Table 5.** Barriers and facilitators to medical device registration.

| Barriers | Specific challenges | Facilitators | Areas for improvement |
| --- | --- | --- | --- |
| Complexity of regulatory rules | Particularly challenging for startups and SMEs as they may lack the experience/in-house examples needed to facilitate independent completion, costs due to the requirement of experienced consultants <sup>23,24,28</sup> | No linked facilitators | Development of templates and examples that can be made freely available to stakeholders on approval by notified bodies <sup>23</sup> |
| Lack of clarity around what constitutes a medical device | A developer's subjective statement about intended use contributes to the determination of whether something is a medical device [18,22]; so, developers could target their device as a lifestyle/well-being app and thus avoid the "medical device" classification [22] | Detailed guidance and documentation available to support the registration process <sup>24,27,28,30,31</sup><br>Specifically, compared to other jurisdictions, the UK provides some of the best guidance on what a software application medical device is a via MHRA <sup>28,31</sup>, NICE (ESF) <sup>27,30</sup>, and MEDDEV guidance documents <sup>28</sup><br>MHRA lists keywords that, if used to describe an app, indicate a need for regulation <sup>28,31</sup><br>MHRA launches an online Eligibility Checker tool for marketing authorisation applications via the new International Recognition Procedure <sup>32</sup> | International consensus on what is considered a 'medical device' to improve clarity around definitions <sup>23,25</sup> |
| Lack of clarity around clinical validation | • Lack of clarity and standardisation around what is considered a significant change and what clinical validation is sufficient for the device <sup>23</sup> | • The NICE ESF (2019) has improved clarity of the evidence required; updated in 2022 to incorporate AI and algorithmic technologies <sup>27,33</sup><br>• NICE guidance provides a framework for the degree of evidence and financial impact required for digital health technologies <sup>26</sup> | International consensus on ‘significant change’ and ‘sufficient clinical evidence’ to improve clarity around validation <sup>23,25</sup> |
| Lack of tailoring to digital health technologies | • Clinical evidence requirements are not tailored to digital health technologies and the potential to use real-world evidence is unacknowledged <sup>23,29–31</sup><br>• Mobile health apps undergo iterative and adaptive development and can make registration and re-review challenging <sup>31</sup><br>• Standard study designs are also generally not suitable as efficacy depends on use in real-world context and not clinical conditions <sup>29,31</sup> | • Different levels of classification can facilitate the process<br>• The relatively simple self-certification process for low-risk (Class 1) medical devices means that most mhealth apps do not need to be certified by a Notified Body <sup>24,26,28</sup><br>• Lower-risk medical devices require less rigorous clinical investigations to evidence efficacy and safety compared to higher-tier devices, facilitating the development of effective, secure apps in line with self-certification policy <sup>26,27</sup> | • Increasing capacity of Notified Bodies by <sup>23</sup>:<br>1. Tailoring specific notified bodies to focus on different tiers of classification<br>2. Offering more training in medical device certification<br>3. Facilitating the process of becoming a notified body<br>• Adapting the registration process to account for the unique characteristics (development and evidentiary) of digital health technologies <sup>24–31</sup><br>• Greater flexibility in evidence generation, suiting the agile, iterative development of digital health technologies <sup>23,27,30,31</sup> |

The lack of clarity on what constitutes a “significant change” in design or intended purpose of a device under MDR Article 120(3) continues to cause confusion in the registration process, despite the existing guidance on significant changes (MDCG 2020-3 ^34^), leading to inconsistent interpretations ^23. 23^. Another common barrier related to the clinical evidence requirements that emerged was that they are not tailored to the specific characteristics of digital health technologies ^23,29–31^. Randomised controlled trials can be difficult to conduct with digital health technologies ^29^, but the potential to use real-world evidence for clinical validation is often unacknowledged ^23^. Real-world evidence refers to “clinical evidence on a medical product’s safety and efficacy that is generated using real-world data resulting from routine healthcare delivery” ^35^. Ongoing development following mobile health app iteration can make registration re-review and approval challenging ^31^. The efficacy of mobile, home-based digital health technologies also depends heavily on how they are used in a real-world context rather than within healthcare environments and clinical evaluations ^29,31^. This affects the suitability of digital health interventions for the standard study designs required for clinical evidence for medical device registration. However, this can be addressed by undertaking clinical validation in the real-world context and environment of use ^29,30^.

### Facilitators to medical device registration for software

Despite barriers in the registration process, several studies highlighted available guidance, including MHRA, NICE (ESF), and MEDDEV documents ^24,27,28,30,31^ - although the purpose of one of these studies was to provide real-world examples to “*help lift the fog of confusion over which regulatory processes apply*” ^24^. The UK, through the ESF, offers strong support for software as a medical device ^27,30^ and NICE provides a framework for evidence and financial impact^26^.

While clinical evidence requirements are not tailored to digital health, different classification levels can aid registration. The self-certification process for low-risk (Class 1) devices simplifies approval, as most mobile health apps do not require Notified Body certification ^24,26,28^. Lower-risk devices also face fewer clinical investigation requirements ^27^, providing motivation for developers to build effective and secure apps in line with self-certification policies ^26^.

### Suggestions for improvement for medical device registration for software

Some studies highlighted potential ways of improving the process. To address clarity issues, achieving international consensus on definitions (e.g., ‘medical device,’ ‘significant change,’ and ‘sufficient clinical evidence’) was suggested ^23,25^. One study proposed developing freely available, notified body-approved templates and examples to streamline the processes ^23^.

Most recommendations focused on adapting regulations to the iterative nature of digital health technologies. Half of the studies emphasized greater flexibility in evidence generation to align with agile development ^23,27,30,31^, as, unlike many medical devices, software as a medical device undergoes regular adaptation and improvement ^31^.

## Discussion

### Summary of findings

Similar themes regarding the barriers and facilitators to the UK medical device registration process emerged from the included papers. The most common barriers were a lack of clarity and specificity in the regulatory rules and definitions that determine whether a digital technology is considered a medical device, what tier it is, and what evidence is sufficient for clinical validation. Several studies also noted that the medical device registration process was not designed to account for specific characteristics of digital health software - such as its ongoing development and the importance of factors that affect real world use - which posed a barrier for clinical evidence gathering and ongoing regulatory compliance. Other barriers included the time-consuming process of registration, high financial costs associated with it, and a lack of access to support, particularly for startups and SMEs.

Interestingly, although fewer facilitators were mentioned, they related to similar issues. Studies reported that guidance documentation was available, and the tiered classification system facilitated the medical device registration process for lower-risk software. Despite the availability of guidance, the barriers and suggestions for improvement identified demonstrate that there is still not sufficient clarity and specificity for stakeholders, who often find the process complex and confusing ^23^. Templates and real-life examples would help to improve clarity and streamline the process. Additionally, it is recommended to provide specific guidance fordifferent types of study designs that can capture real world evidence for clinical validation.

### Strengths and weaknesses of the studies

Only two of the studies ^23,30^ gathered opinions from key stakeholders involved in medical device registration (including innovators and developers, healthcare and compliance consultants, notified bodies, policy makers, health authorities, patient associations, and others). The remaining three quarters of the studies (6/8) were perspective papers or case studies that presented the authors’ interpretation of the barriers and facilitators to the UK’s software as a medical device registration process ^24,25,27–29,31^. While still useful, this potentially biases the results of this review towards a more academic stakeholder perspective of the barriers and challenges to UK medical device registration. There was general alignment between the themes identified in the qualitative study with stakeholders ^23^ and those discussed in the other studies, which supports the findings of that study.

### Strengths and limitations of the review

One of the limitations of the review is that the retrieved references were primarily academic articles. The search initially included Google Scholar to capture more of the grey literature, which might have included more resources discussing barriers and facilitators to the UK medical device registration process. The search identified 1,520 references, but it was not possible to export them all together, and an attempt to download them individually resulted in Google Scholar limiting the amount of references that could be exported to about 100. Due to time constraints, this meant that Google Scholar had to be removed from the search. The search terms did not include “MDD” which could have potentially missed some relevant results; however, it is likely that all relevant papers would be captured with the terms “medical device” or “CE mark” (the output of the MDD), which were included. Additional medical device terms were identified from the studies retrieved (such as IMDRF); these may be useful to include in future reviews but were not specific to the scope of this review, which focused on a UK context. Eligibility criteria used the year of smartphone availability (2008) as a cut-off for inclusion; this could have biased results towards patient-rather than clinician-facing technologies, as web-based decision support tools were in use prior to this. This decision reflected an aim to examine the recent landscape of device registration, but cut-offs in future research might be better to focus on changes in the MDR landscape instead. The search and screening of literature were conducted from October to November 2022. Given the fast advances in healthcare policies and digital technologies, findings should be interpreted with an awareness that the landscape may have changed since the search was undertaken. To ensure relevance, we attempted to refer to the latest publicly available policies at the time of writing. Subsequent researchers are suggested to conduct an updated review to capture the most current evidence. Another limitation is that the title and abstract screening was only conducted by one author. To improve the transparency of the review, the PRISMA-ScR framework ^21^ was used to ensure that all of the requirements for a scoping review were conducted and two authors independently screened the full texts to determine final inclusion. The potential bias in the initial screening of the references means that there is a possibility that some relevant papers may have been missed. Given the available resources and time, it was not possible for both reviewers to independently conduct the full screening process.

### Meaning and future research

The findings from this study highlight the importance of updating medical device regulation to address the specifics of digital health technologies. Although the studies included in this review are relatively recent (published between 2015 and 2022), a recent MHRA consultation with stakeholders identified similar issues and suggestions to those captured in this review, including improving the clarity around definitions and guidance and aligning definitions and risk classifications with international standards and other regulatory bodies ^16^. One issue may be the lack of familiarity of developers with the healthcare landscape, a potential means of addressing this would involve building relationships and shared professional understanding of the context of use, or clinical trial expertise within the companies developing the technologies, particularly for SMEs.

The barriers associated with registering software as a medical device have the potential to impact the way that digital health technologies are promoted, adopted, and used. This is particularly true in the grey areas around the registration; previous studies have found that there are numerous mobile health apps that are publically available that, based on their stated intended use, should be classified as medical devices but are not ^5,36^. The lack of clarity in the definitions of medical devices likely contributes to this. Although the digital health technologies that are in these grey areas are likely to be lower risk, it has potential implications for patient safety. As much of this grey area hinges on developer-stated intended uses and audiences, it would be interesting for future research to investigate the degree of alignment between intended and actual use of digital health technologies. There is also further potential for confusion as released apps are updated and developed further; what originally may not have been classified as a medical device could become one as it evolves or it may need a stricter level of regulation, and would require a reclassification process to be conducted ^28^.

The current structure of the software as a medical device registration process also has potential implications for innovation in digital health technology. There is a delicate balance between the slow process of regulation to ensure safety and the rapid innovation processes in digital health ^25^. Too permissive a regulatory system, without a stringent assessment of clinical evidence, can enable software to be certified without sufficient safety evidence; a basis which could then be used to certify similar devices that are ‘substantially equivalent’ ^13^. On the other hand, excessive regulatory bureaucracy can hinder innovation and reduce incentives to adopt and improve new technologies ^37^. The current registration process has been criticised for inhibiting innovation and development due to its rigid and lengthy processes which do not reflect the nature of digital health and the ever changing landscape of technologies ^23^. Often, developers do not have the time or resources for the current regulation process, as their product will have developed into a new stage or version by the time the process is complete ^31^. One study found a facilitator was clinical engagement, and funders wanting to see collaborative bids provides an incentive for developers to seek clinical engagement. The barrier now is lack of expertise and time to engage within clinical and academic organisations, which will require a separate set of policies and solutions.

To help address the concern that regulation might be stifling digital innovation and to adapt the regulation process to better fit the realities of digital health technology development, the FDA recently introduced a pilot pre-certification (“Pre-Cert”) program ^13,38^. This program shifts towards the use of post-market, real-world evidence evaluations and enables streamlined review for software from companies that have been pre-certified ^38^, but it still needs greater transparency and detail in the real-world metrics and adverse event reporting used to assess the technologies ^13^. The shift towards real-world evidence has potential, as the clinical evidence that is commonly used for proving effectiveness, accuracy and so on, is not always relevant and sometimes can be very difficult to apply to digital health technologies, with the use of real world data being often overlooked within this domain ^23,29^. Potential issues with real world data are the self-selecting nature of the patients who use the apps, the non-standardised collection of data, lack of fidelity in intervention implementation, the lack of robust outcomes, and the inability to adjust for confounders due to lack of linkage with routine health and care datasets. Further research should focus on identifying ways in which to investigate the effectiveness of digital health technologies, with particular focus on translating real world data to ensure the technologies are fit for purpose and user interface and at what point in the process this evidence would be more appropriate (for example, initial approval processes vs. on-going safety monitoring).

## Conclusion

This scoping review aimed to explore and summarise the key barriers and facilitators to software as a medical device registration in the UK. The main issues that were identified revolved around a lack of clarity and specificity in definitions and guidance for the registration and regulation of digital health technologies and a lack of tailoring of those regulations to the specific characteristics of digital health technologies. This is a topical review, as the UK’s MHRA recently updated its guidance on software and artificial intelligence as medical devices on 3 February 2025. Several of the papers included suggestions for how these barriers could be addressed, to the benefit of the stakeholders involved in the medical device registration process as well as the patients and healthcare professionals who are the end users of the digital health technologies. For digital health to fully flourish, it is important to ensure the processes reflect the demand for development and allow for technologies to continually adapt with the evidence, patient and clinician need, and societal climate. There is a need to ensure that the regulation process is formulated to align with agile methods of development and the critical need for validation and safety. This could enable more efficient, continuous innovation of technologies without burdensome processes, ensuring that users receive the most safe and effective tools possible. Further research could examine how barriers in the medical device registration process affect these end users and which suggestions have the greatest potential benefit for the ease and efficiency of regulation and the safety and positive impact for patients and healthcare professionals.

## Data Availability

The authors confirm that the data supporting the findings of this study are available within the article and its supplementary materials.

## Acknowledgments

The review was conceived and supervised by EM. MMI conducted the searches, screening, data extraction, and analysis and drafted the first version of the review. KB conducted full text screening and analysis and contributed to the drafting of the manuscript. All authors contributed revisions, with final revisions by MMI, CC and EM.

## Supporting information

S1 Appendix. PRISMA-ScR Checklist

S2 Appendix. Search record

S3 Appendix. Endnote search criteria

